# Development and Validation of CPX-MATE: An End-to-End Medical Education Platform Integrating Voice-Based Virtual Patient Simulation and Automated Real-time Evaluation

**DOI:** 10.64898/2026.02.21.26346803

**Authors:** Ji Woo Song, Minseong Kim, Chanhee Hong, Young Sam Kim, Junho Cho, Ji Hoon Kim, Jinwoo Myung, Arom Choi, Hanna Yoon, Stephen Gyung Won Lee, Seng Chan You, Chaeryoung Park

## Abstract

**Background:** Objective Structured Clinical Examination (OSCE; Clinical Performance Examination [CPX] in South Korea) is a high-stakes assessment of clinical performance, communication, and reasoning during time-limited patient encounters. As AI-enabled virtual standardized patient (VSP) simulation and automated scoring are introduced for OSCE-like training, prospective evidence is needed on how such systems perform and are perceived when embedded in real educational workflows.

**Methods:** We developed CPX with Medical students’ Assistant for Training and Evaluation (CPX-MATE), a web-based platform integrating (1) CPX with Virtual Standardized Patient (CPX-VSP), real-time voice dialogue with a VSP using speech-to-speech (STS) models, and (2) CPX with Real-Time Evaluator (CPX-RTE), automated transcription, checklist-based scoring, and feedback from encounter audio using a Speech-to-Text model and a large language model. During an emergency medicine clerkship (Nov 2025-Jan 2026), 60 senior medical students completed two 12-min CPX encounters (VSP with acute pancreatitis; HSP with ureteral stone) with immediate CPX-RTE feedback. For CPX-VSP, students were assigned to either a full-capacity or a resource-limited STS configuration (n=30 each). Dialogue fidelity was evaluated by turn-by-turn analysis of student–VSP exchanges, classifying responses into clinically meaningful error types (tangential, oversharing, role-breaking, off-script). CPX-RTE performance was assessed by agreement (Gwet’s AC1) with professor real-time and resident video-based ratings using a 45-item checklist. Usability of CPX-VSP and CPX-RTE, with overall system usability scale (SUS), were surveyed, and mean per-session costs for CPX-VSP and CPX-RTE were calculated.

**Results:** Across 3,282 dialogue turns, overall error rates were 1.77% versus 9.43% for full-capacity versus resource-limited STS configurations (p<0.001), driven by fewer tangential and oversharing responses; no off-script errors were observed. The mean per-session cost was $0.12 for resource-limited configuration and $0.78 for full-capacity configuration. CPX-RTE showed high agreement with human ratings (AC1=0.916 vs professor; 0.916 vs resident), with slightly different levels of agreement across four sections, and high usability across all domains (mean scores, 4.65–4.92), with a per-session cost of $0.17. CPX-MATE demonstrated good overall usability (median [IQR] of 77.5 [70.0-85.0]).

**Conclusions:** Embedded within a prospective clinical clerkship, CPX-MATE demonstrated operational fidelity and human-level checklist agreement as an end-to-end, voice-based AI-assisted OSCE platform. This real-world deployment supports its scalable integration as a complementary assessment tool while highlighting the importance of systematic validation and context-aware implementation in medical education.

## INTRODUCTION

Objective Structured Clinical Examination (OSCE), which is called Clinical Performance Examination (CPX) in South Korea, is a core competency-based assessment designed to evaluate authentic clinical performance of medical students through standardized patient (SP) encounters. ^1,2^ OSCE is a high-stakes component of undergraduate medical education and licensure, assessing integrated skills such as history taking, communication, clinical reasoning, and initial management within time-limited, face-to-face interactions. Students interact directly with an SP, while evaluators observe in real time from behind a one-way mirror and score student performance using standardized checklists.

Despite its strong educational validity, the educational infrastructure underpinning OSCE is inherently resource intensive. It requires substantial financial investment, complex logistical coordination, trained faculty examiners, and the recruitment, training, and compensation of human standardized patients (HSPs).^3,4^ These demands limit opportunities for repeated practice, timely feedback, and scalable deployment. Such challenges are particularly pronounced in low- and middle-income countries (LMICs), where constrained educational budgets, faculty shortages, and limited access to trained SPs hinder the sustainable delivery of OSCE-based education.^5,6^

Recent advances in artificial intelligence (AI), particularly large language models (LLMs) and speech-to-speech (STS) models, have generated growing interest in virtual standardized patient (VSP)-based simulations as a means to augment traditional OSCE education. Prior studies have demonstrated the technical feasibility of text-based VSPs and AI-assisted automated scoring systems.^3,7-16^ However, much of the existing literature has focused on whether such systems can be built, rather than on how they should be systematically evaluated and integrated into real-world medical education. In high-stakes clinical assessment, technical performance alone is insufficient; perceived realism, learner trust, and sustained usability are equally critical determinants of educational impact. At the same time, reliance on high-capacity models raises questions about accessibility and cost equity.

Another key limitation of prior AI-based OSCE solutions lies in feedback delivery. Although automated scoring modules have been developed, most were designed for retrospective evaluation rather than for real-time, formative feedback during ongoing practice.^17-19^ Timely feedback is a core principle of experiential learning, enabling learners to identify performance gaps and adjust clinical reasoning while the learning experience remains cognitively salient.

Accordingly, we developed and evaluated CPX with Medical students’ Assistant for Training and Evaluation (CPX-MATE), an end-to-end platform integrating voice-based VSP training (CPX with Virtual Standardized Patient, CPX-VSP) and real-time, voice-based evaluation applicable to HSP encounters (CPX with Real-Time Evaluator, CPX-RTE). This study aims to define and empirically examine the dimensions that should be scrutinized as such systems enter routine educational use, with a particular focus on performance fidelity, usability, and deployment within authentic educational settings.

## METHODS

### Development of CPX-MATE

#### System Overview

CPX-MATE is a web-based system comprising CPX-VSP and CPX-RTE (Figure 1). CPX-VSP enables real-time voice dialogue with a VSP using two STS models (*gpt-realtime* and *gpt-realtime-mini*).^14^ This dual-model design was adopted to examine whether a resource-limited configuration can provide acceptable performance in constrained settings within the rapidly evolving AI landscape, and to quantify the extent of performance gains achievable with a full-capacity model as AI capabilities advance. To simplify presentation, *gpt-realtime* is hereafter referred to as the full-capacity model, whereas *gpt-realtime-mini* is referred to as the resource-limited model. CPX-RTE transcribes HSP encounters using speech-to-text (STT) model (*whisper-1*)^20^, maps transcript evidence to predefined checklist items and generates feedback using an LLM (*GPT-5*)^21^.

**Figure 1.**
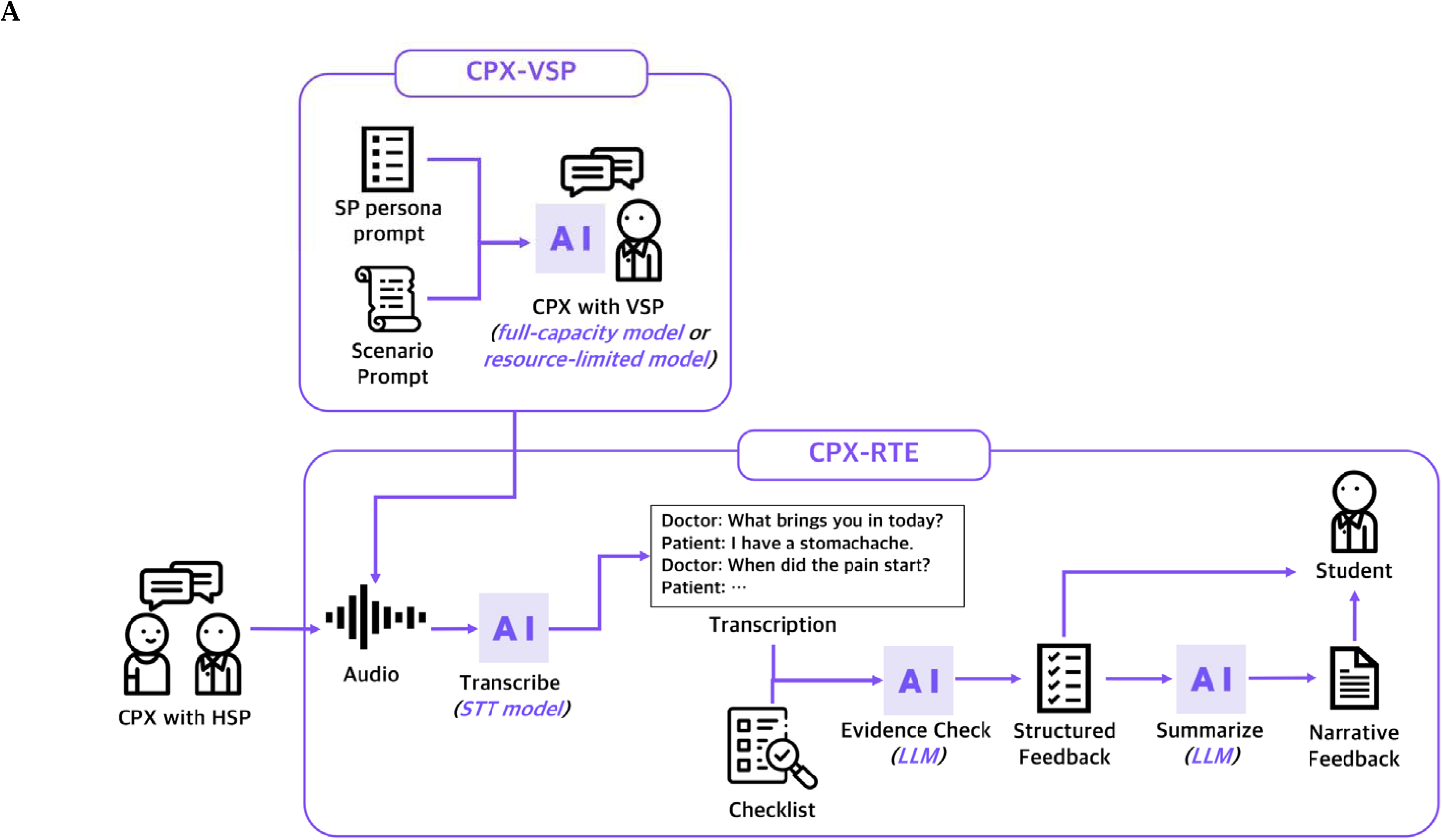
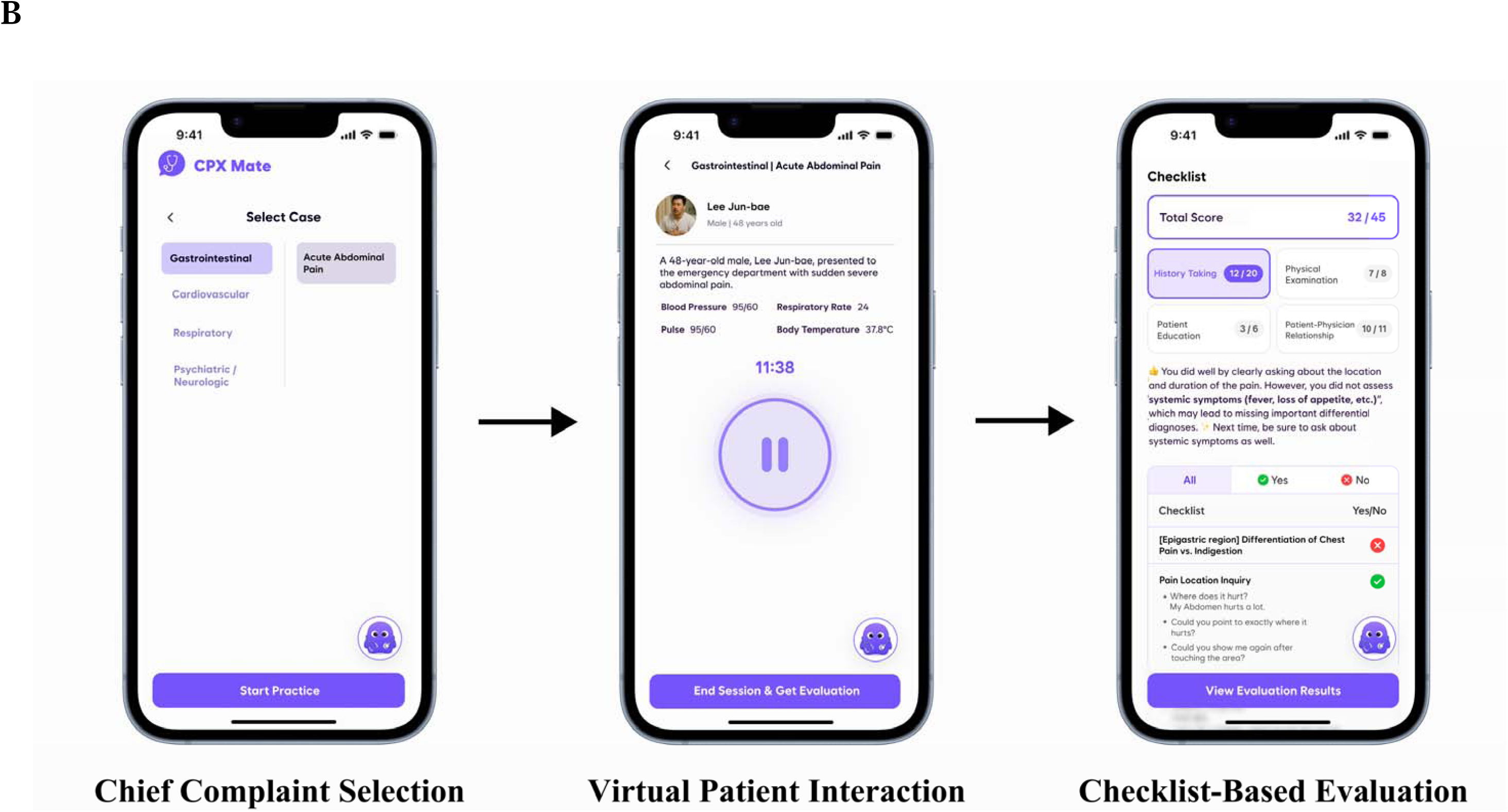
Overview of the CPX-Mate Platform. (A) System architecture of CPX-Mate for CPX with VSP and CPX with Real-Time Evaluator which contains virtual standardized patients simulation using STS (speech-t speech) model and automatic, checklist-based assessment using STT (speech-to-text) model large language models (LLMs). (B) Mobile Web-app workflow of CPX-Mate Users select a chief complaint, interact with a virtual patient through real-time dialogue (CPX-VSP), and receive checklist-based performance evaluation and feedback (C RTE).

#### CPX-VSP: prompts and interaction rules

Two prompts were provided to the STS model: (1) a global SP persona prompt, which was identical across all chief complaints and scenarios, specifying role/tone/constraints and dialogue rules (e.g., disclose information only when asked; answer unspecified questions plausibly without contradicting the scenario; if the student announces a physical exam, respond with corresponding findings), and (2) a scenario-specific prompt which varied by clinical scenario and contained the full clinical case specification (symptoms, history, and physical exam findings). The SP persona prompt was designed as a reusable, chief-complaint–agnostic control layer, whereas the scenario prompt served as a case-specific content layer. Although only a single scenario prompt was used in the present study, this separation was intentionally adopted to ensure scalability to multiple chief complaints and scenarios. Prompts were iteratively refined by an emergency medicine faculty member (CP) and a medical student investigator (JWS).

#### CPX-RTE: scoring and feedback

CPX-RTE follows a pipeline consisting of (1) audio transcription, (2) structured checklist-based feedback generation, and (3) narrative feedback generation. The student–patient dialogue is first transcribed using an STT model. For each checklist item, an LLM then reviews the transcript to identify utterances that match a criterion of the item and applies a predefined binary decision rule, classifying the item as “Yes” when one or more relevant utterances are detected and as “No” otherwise. These item-level decisions, together with the matched transcript excerpts, form the structured feedback. Finally, the LLM generates narrative feedback summarizing strengths and areas for improvement based on both the transcript and the structured outputs.

#### Clinical scenarios and checklist

Two acute abdominal pain presentations were used as representative, generalizable emergency cases: VSP (CPX-VSP) acute pancreatitis and HSP ureteral stone. Performance was evaluated using a 45-item acute abdominal pain checklist: History Taking (20), Physical Examination (8), Patient Education (6), and Patient–Physician Interaction (11), developed with reference to the Korea Association of Medical Colleges’ The guide to clinical performance, 2nd edition.^22^

### Validation of CPX-MATE

#### Study design, setting, and participants

The study was conducted during an emergency medicine clerkship at Yonsei University College of Medicine between November 2025 and January 2026 and involved year 3–4 medical students. Each participant completed a standardized CPX experience consisting of two clinical encounters followed by a post-session survey.

Participants first engaged in a 12-minute CPX encounter with a VSP, during which CPX-VSP was delivered using either a resource-limited STS model (n=30, first half of the study period) or a full-capacity STS model (n=30, second half). CPX-RTE generated real-time checklist-based feedback that was shown to students for formative learning during this encounter, but these outputs were not used for performance validation as no independent human scoring was performed for VSP encounters.

After a 5-minute intermission, participants completed a second 12-minute CPX encounter with an HSP. These HSP encounters were audio-recorded for automated scoring by CPX-RTE, while video recording was performed as part of routine CPX operations to enable independent human rating. HSP performance was scored in real time by a board-certified emergency medicine attending physician (CP) and later independently scored by an emergency medicine resident (CH) using the same 45-item checklist. CPX-RTE scoring results were also generated and presented to students immediately after the encounter.

Following completion of both encounters, participants completed post-session surveys assessing the usability and perceived realism of CPX-VSP, the quality of CPX-RTE feedback, and overall system usability, along with open-ended questions for qualitative feedback.

#### Data collection

Collected data included CPX-VSP transcripts and system logs (e.g., turns, duration, token usage), HSP audio (and video for human rating), and post-session surveys (Likert-scale items and free-text responses). All data were de-identified before analysis. Mean per-session costs for CPX-VSP and CPX-RTE were captured through OpenAI’s dashboard.

#### Outcome Measures

CPX-VSP performance was assessed at the Minimum Interaction Unit (MIU) level, defined as one student utterance paired with the corresponding VSP response. Each MIU was classified into one of four error types: tangential (plausible content misaligned with question intent), oversharing (unsolicited disclosure beyond what was asked), role-breaking (the VSP speaking as if it were the clinician), and off-script (responses contradicting the predefined scenario prompt). All MIUs were manually reviewed and coded by a medical student investigator (JWS). The primary performance metric was the overall error rate per MIU for each STS model; secondary metrics were error rates per MIU stratified by error type.

CPX-VSP usability was assessed using post-session surveys adapted from Cook et al. on a 1-to-6 Likert scale (1 = strongly disagree, 6 = strongly agree). Survey items evaluated two domains: (1) dialogue naturalness (Humanlike, Coherent, Clinically relevant, Dialogue overall) and (2) user experience(UX)/immersion (Realness, Cognitive authenticity, Variability, Involvement, UX overall). One Cook et al. item assessing whether the patient’s “personal preferences” were reflected was excluded to preserve construct alignment with the target assessment context. In the Korean undergraduate CPX setting, checklist-based scoring typically emphasizes standardized history taking, clinical reasoning, and initial management/education under time constraints rather than individualized preference elicitation as a primary learning objective. Accordingly, we focused the usability instrument on dialogue naturalness constructs most relevant to CPX preparation.

CPX-RTE performance was evaluated using Gwet’s AC1 for binary checklist items. AC1 was used since checklist items showed marked prevalence imbalance (some items were almost always performed, whereas others were rarely performed), a setting in which kappa-type coefficients can be unstable and underestimate agreement. AC1 is well-known for providing a robust agreement estimate under such imbalanced marginal distributions.

The board certified emergency medicine attending physician (CP) scored the encounter in real time using the same 45-item acute abdominal pain checklist, while a second rater (CH, emergency medicine resident) independently scored the video-recorded encounter. Agreement was evaluated across three rater pairs: CPX-RTE vs attending, CPX-RTE vs resident, and attending vs resident. The primary outcome was overall agreement across all 45 checklist items for each pair, with secondary analyses examining section-level agreement (History Taking, Physical Examination, Patient Education, and Patient–Physician Interaction) and item-level agreement for each checklist item.

CPX-RTE usability was assessed using post-session surveys adapted from Cook et al. on a 1-to-6 Likert scale across four constructs: Evidence-based, Actionable, Connected, and Balanced.

Participants were asked whether they observed biased or stereotyped content (e.g., related to sex, age, race/ethnicity, or socioeconomic status) in either CPX-VSP dialogue or CPX-RTE feedback, and to describe the bias if present. Overall web application usability was assessed using the System Usability Scale (SUS).^23^

To promote objective and consistent responses, surveys assessing CPX-VSP realism and user experience and CPX-RTE feedback quality were administered using 1–6 Likert scales accompanied by explicit operational criteria defining each response level. Participants also answered open-ended questions on factors that enhanced or hindered perceived authenticity of CPX-VSP, and on areas for improvement of CPX-RTE. These free-text responses were analyzed using inductive thematic analysis framework suggested by Braun and Clarke.^24^The verbatim survey items are provided in <u>Supplementary Method 1</u>.

#### Statistical analysis

Analyses were conducted using a complete-case approach. For CPX-VSP performance, errors were defined at the MIU level. Overall error rates and error rates by subtype were calculated per STS model. CPX-VSP usability was summarized as mean (95% confidence interval [CI]) and full frequency distributions (n, %). Participant-level cluster bootstrap resampling (1,000 iterations) was used to obtain 95% CIs and p-values. Since CPX-VSP performance and usability analysis compared two difference STS models, all multiple comparisons for CPX-VSP were adjusted using the Benjamini–Hochberg false discovery rate procedure. All tests were two-sided with a significance threshold of α=0.05.

For CPX-RTE performance, agreement on binary checklist items was assessed using Gwet’s AC1 to account for marginal imbalance. CPX-RTE usability items were reported as mean (95% CI) and full frequency distributions (n, %). Similarly, participant-level cluster bootstrap resampling (1,000 iterations) was used to obtain 95% CIs. Analyses were performed using Python (ver 3.14.2; Python Software Foundation).

#### Ethics

The study was approved by the IRB of Yonsei University College of Medicine (IRB No. [4-2025-1312]). Participation was voluntary, uncompensated, and had no impact on academic evaluation; all recordings/transcripts were de-identified.

## RESULTS

### Participants

Sixty senior medical students (years 3–4) completed the sequential two-station CPX sessions between November 2025 and January 2026. Participants had a mean age of 26.9 years (SD 0.76) and included 28 female (46.7%) and 32 male (53.3%) students. CPX-VSP was delivered using the resource-limited model (n=30) or the full-capacity model (n=30). Detailed demographics are shown in Table 1.

**Table 1.** Participant demographics by model group.

|  | Participants<br>N | Male<br>N (%) | Age, years<br>mean (SD) | 3<sup>rd</sup> grade<br>N (%) |
| --- | --- | --- | --- | --- |
| **Resource-limited**<br>**model** | 30 | 18 (60.0%) | 27.2 (0.85) | 4 (13.3%) |
| **Full-capacity**<br>**model** | 30 | 14 (46.7%) | 26.5 (0.35) | 16 (53.3%) |
| **Total** | 60 | 32 (53.3%) | 26.9 (0.76) | 20 (33.3%) |

Baseline demographic characteristics of participants stratified by model group. Age is presented as mean (standard deviation), and categorical variables are presented as number (percentage).

### Performance of CPX-VSP: Dialogue Error Rates per MIU

Across CPX-VSP sessions, 3,282 Minimum Interaction Units (MIUs) were identified and manually reviewed (resource-limited model: 1,591 MIUs; full-capacity model: 1,691 MIUs). Figure 2 shows the dialogue error rates per MIU of each model. The overall MIU error rate was higher with the resource-limited model than with the full-capacity model (9.43% [95% CI 7.50–11.25] vs 1.77% [0.80–2.84]), yielding an absolute difference of 7.65 percentage points (resource-limited − full-capacity; 95% CI 5.34–9.74; p<0.001) (Fig. 1). Error subtype analyses showed significantly higher tangential and oversharing error rates for the resource-limited model (tangential 5.28% [3.97–6.69] vs 1.48% [0.66–2.40], Δ=3.80 pp [2.23–5.40], p<0.001; oversharing 4.02% [2.86–5.33] vs 0.24% [0.06–0.46], Δ=3.79 pp [2.60–5.09], p<0.001). Role-breaking was rare in both groups (0.13% vs 0.06%, Δ=0.07 pp [−0.12 to 0.27], p=0.485), and off-script errors were not observed.

**Figure 2.**
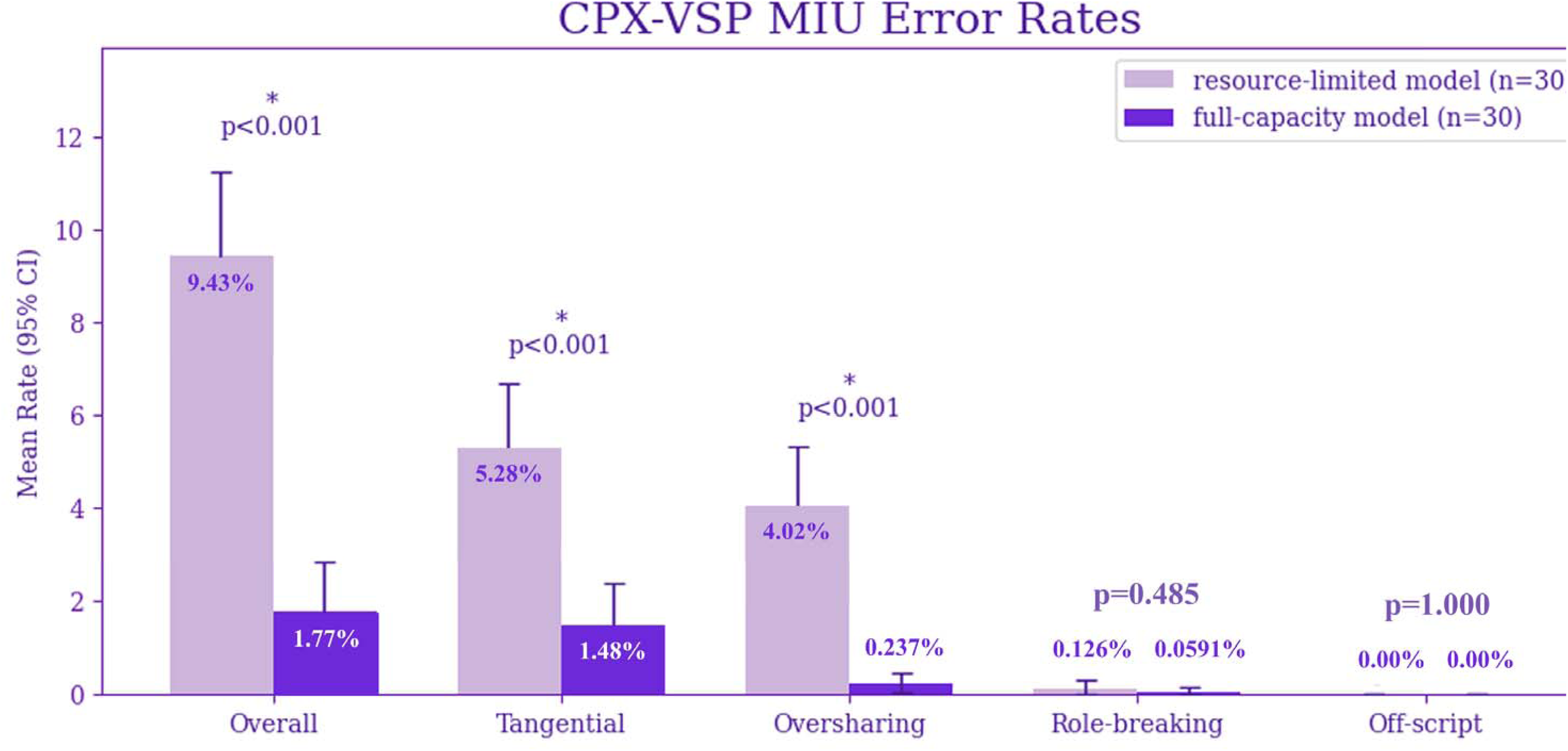
CPX-VSP MIU error rates across error subtypes. Tangential answers refer to responses that are factually correct but misaligned with the student’s question intent; oversharing answers disclose unsolicited information bey what was asked; off-script answers contradict the predefined scenario prompts.

### Usability of CPX-VSP: Dialogue Naturalness and User Experience

As shown in Figure 3, the full-capacity model received higher ratings than the resource-limited model for Coherent (4.77 [95% CI 4.43–5.13] vs 4.03 [3.67–4.43]; p=0.034), Involvement (4.37 [4.03–4.70] vs 3.40 [2.87–3.90]; p=0.019), and UX overall (4.27 [3.90–4.60] vs 3.27 [2.83–3.63]; p=0.010). While differences in other usability items did not reach statistical significance, ratings consistently favored the full-capacity model over the resource-limited model across all measures. Detailed results are shown in Supplementary Table S1.

**Figure 3.**
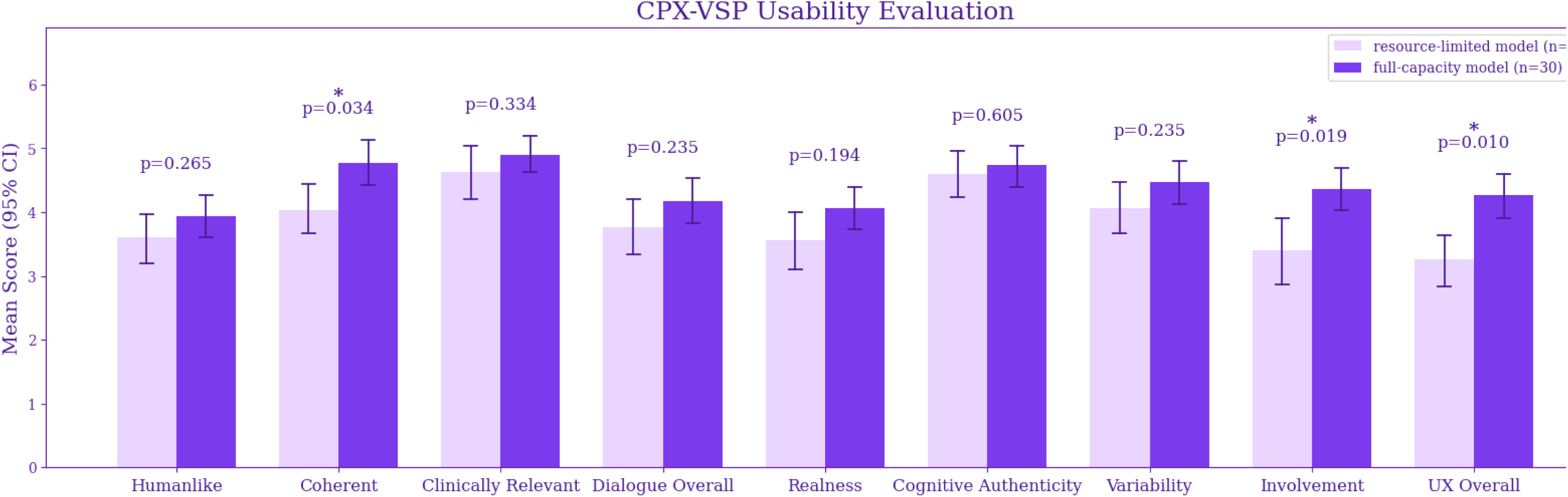
Comparison of CPX-VSP Usability and User Experience Scores Between Models. *Humanlike*, *Coherent*, *Clinically Relevant*, and *Dialogue Overall* assess the naturalness, consistency, clinical usefulness, and overall realism of the dialogue, respectively, while *Realness*, *Cognitive Authenticity*, *Variability*, *Involvement*, and *UX Overall* evaluate realism, cognitive engagement, spontaneity, immersion, and overall user exper respectively.

Open-ended responses aligned with quantitative findings. Students most frequently attributed authenticity to content-driven immersion, particularly coherent, clinically plausible, and scenario-consistent responses (Supplementary Table S2). Loss of immersion was primarily linked to content-level inconsistencies, including tangential responses and oversharing (Supplementary Table S3). Students also emphasized interactional realism (e.g., patient-like tone and affective responsiveness), consistent with higher involvement and overall UX in the full-model group. Once major content- and interaction-level issues were reduced in the full-capacity model, secondary constraints (e.g., latency and screen-mediated awkwardness) became more prominent in qualitative comments.

### Performance of CPX-RTE: Agreement with Human Ratings

Checklist scoring by CPX-RTE was compared with professor real-time ratings and resident video-based ratings using Gwet’s AC1 (Table 2). Overall agreement was 0.916 (95% CI 0.902–0.930) for CPX-RTE vs professor, 0.916 (0.903–0.929) for CPX-RTE vs resident, and 0.976 (0.968–0.983) for professor vs resident. Section-level agreement between CPX-RTE and human evaluator was highest for History Taking (CPX-RTE vs professor 0.957 [95% CI 0.944–0.971]; CPX-RTE vs resident 0.958 [0.943–0.972]) and lowest for Patient–Physician Interaction (CPX-RTE vs professor 0.860 [0.823–0.893]; CPX-RTE vs resident 0.847, [0.809–0.881])

**Table 2.** Inter-rater Agreement (Gwet’s AC1) Between CPX-RTE and Human Raters by Checklist Section.

| Section | Gwet's AC1 (95% CI) |  |  |
| --- | --- | --- | --- |
|  | CPX-RTE vs<br>Attending | CPX-RTE vs<br>Resident | Attending vs Resident |
| Overall | 0.916 (0.902–0.930) | 0.916 (0.903–0.929) | 0.976 (0.968–0.983) |
| History | 0.957 (0.944–0.971) | 0.958 (0.943–0.972) | 0.983 (0.974–0.991) |
| Physical Exam | 0.905 (0.870–0.935) | 0.912 (0.878–0.943) | 0.982 (0.966–0.994) |
| Patient Education | 0.906 (0.866–0.944) | 0.921 (0.881–0.956) | 0.965 (0.934–0.990) |
| Patient-Physician<br>Interaction | 0.860 (0.823–0.893) | 0.847 (0.809–0.881) | 0.963 (0.945–0.979) |

The checklist consisted of four sections: History (20 items), Physical Examination (8 items), Patient Education (6 items), and Patient–Physician Interaction (11 items). Gwet's AC1 was computed at the case level within each section using checklist items as units of agreement, and section-level summary statistics (mean [95% CI]) are shown.

Item-level analyses revealed that the three checklist items with the lowest agreement consistently occurred within the Patient–Physician Interaction domain for both CPX-RTE–professor and CPX-RTE–resident comparisons (Supplementary Table S4). Among these, the item “Explore patient’s deep concern” demonstrated the poorest agreement, with an AC1 of 0.250 for both CPX-RTE versus professor and CPX-RTE versus resident, whereas agreement between the resident and professor raters for the same item remained high (AC1 = 0.937).

### Usability of CPX-RTE: Feedback Quality and Refinement Suggestions

60 Participants rated CPX-RTE feedback quality (Evidence-based, Actionable, Connected, Balanced) on 1–6 Likert scales. The overall CPX-RTE feedback quality score (mean of the four items) was 4.82 [4.65–4.99] out of 6, indicating generally high perceived feedback quality; detailed item-level ratings are reported in Table 3.

**Table 3.**
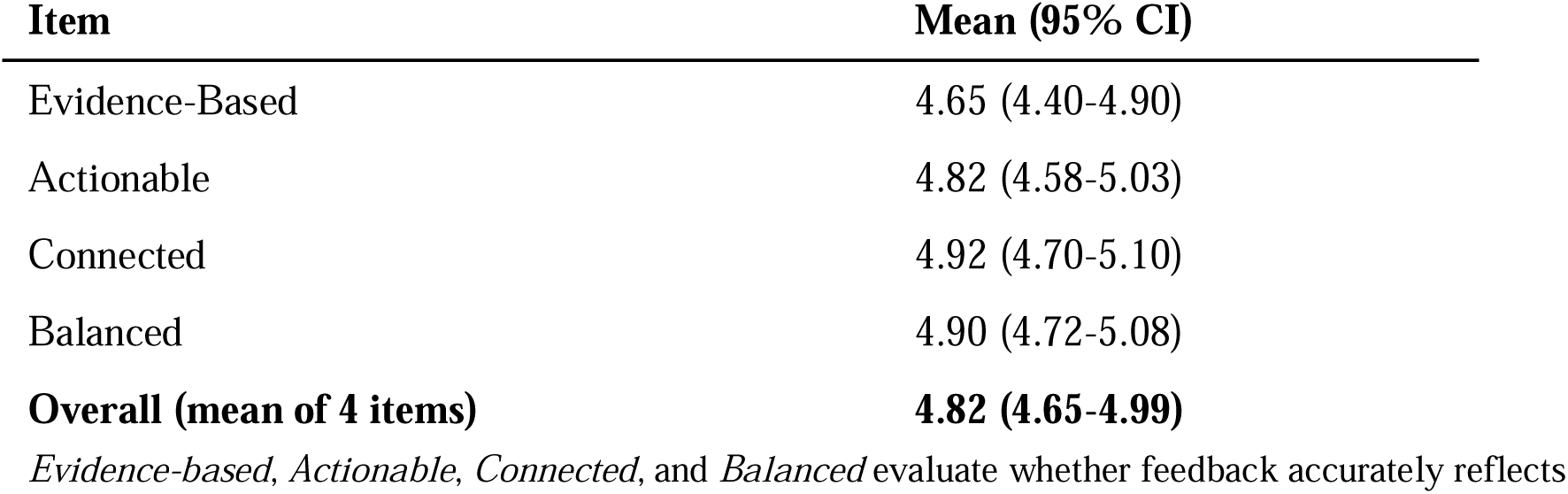
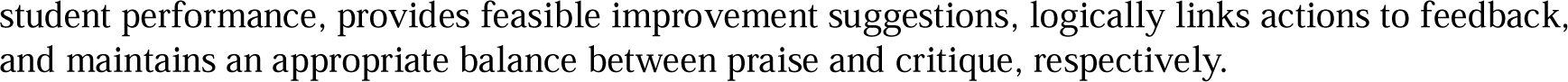
Usability Evaluation of CPX-RTE Feedback.

| Item | Mean (95% CI) |
| --- | --- |
| Evidence-Based | 4.65 (4.40-4.90) |
| Actionable | 4.82 (4.58-5.03) |
| Connected | 4.92 (4.70-5.10) |
| Balanced | 4.90 (4.72-5.08) |
| **Overall (mean of 4 items)** | **4.82 (4.65-4.99)** |

*Evidence-based, Actionable, Connected, and Balanced* evaluate whether feedback accurately reflects
student performance, provides feasible improvement suggestions, logically links actions to feedback, and maintains an appropriate balance between praise and critique, respectively.

For CPX-RTE, open-ended responses highlighted scoring accuracy as the most frequent concern, particularly false-negative detections of performed checklist items (Supplementary Table S5). Despite this, learners consistently reported that the feedback was educationally valuable, emphasizing needs for clearer prioritization, actionable examples, and longitudinal tracking, complementing the high overall usability ratings observed in structured surveys.

### Bias Assessment, System Usability, and Cost

None of the participants reported perceived bias or stereotyped content in CPX-VSP dialogue and/or CPX-RTE feedback. Overall system usability, measured by the System Usability Scale (SUS), had a median score of 77.5 (IQR, 70.0-85.0), indicating generally good usability. The mean per-session cost of CPX-VSP was $0.12 for resource-limited configuration and $0.78 for full-capacity configuration, reflecting a substantial cost–capability differential. A per-session cost of CPX-RTE was $0.17.

## DISCUSSION

This study demonstrates that an end-to-end AI-supported OSCE training loop—voice-based VSP simulation (CPX-VSP) followed by immediate, structured evaluation and feedback (CPX-RTE)—can be deployed within an authentic clerkship workflow with educationally meaningful usability and performance. Across objective and subjective measures, CPX-VSP supported OSCE-oriented practice through sufficiently coherent and immersive dialogue, while CPX-RTE showed high agreement with expert human ratings for checklist-based performance and was perceived by learners as evidence-based, actionable, and balanced. From the learner’s perspective, CPX-MATE functioned as a single continuous cycle—simulation followed by immediate feedback—highlighting the practical premise that OSCE preparation requires not only a realistic encounter but also timely, structured evaluation for deliberate practice.

A key contribution of this work is the use of STS models to achieve real-time, voice-based VSP interaction, addressing a persistent limitation of prior AI patient simulators. Prior studies have demonstrated the educational potential of generative AI–based patient simulators. For example, Lavigne et al. showed that a GPT-4–based AI standardized clinical examination simulator can improve OSCE performance, ^3^ but interaction in that system was entirely text-based, limiting its ability to capture the temporal, prosodic, and turn-taking dynamics that characterize authentic clinical dialogue. Likewise, Luo et al. reported that an LLM-based digital patient system enhanced ophthalmology history-taking skills and incorporated voice interaction via a chained STT–LLM–TTS architecture.^10^ However, learners were required to review and correct transcribed text before eliciting a spoken response, introducing an explicit verification step that interrupts conversational flow. By integrating STS models directly into the voice loop, the present work reduces these sequenced bottlenecks and supports more continuous, contextually responsive spoken interaction.

Furthermore, since STS models are relatively recent and have not been systematically characterized in medical simulation settings, rigorous and fine-grained evaluation was necessary. Accordingly, we decomposed more than three thousand dialogue turns into MIUs and classified failures into four clinically meaningful error types, enabling identification of characteristic instruction-following and conversational control limitations that are invisible to aggregate realism scores alone. This MIU-level validation framework shifts validation from subjective impressions or downstream outcomes to mechanistic understanding of model behavior and provides a transferable methodology for assessing whether emerging STS models are educationally fit for use in high-stakes clinical skills training.

This study also advances prior work on automated OSCE evaluation by prospectively operationalizing real-time, learner-facing assessment within routine clinical education and formally evaluating its usability. Unlike earlier studies that demonstrated feasibility using retrospective or offline transcript analysis,^17-19^ CPX-RTE delivered checklist-based scoring and structured narrative feedback to medical students within seconds after each encounter, at scale, during routine clinical education. Importantly, this real-time feedback was not only technically validated against expert raters but also systematically evaluated by survey of usability from sixty end users, providing direct evidence on how immediate AI-generated feedback is perceived, interpreted, and integrated in real educational practice rather than simulated or post hoc settings.

At the same time, our findings clarify clear boundaries of what automated evaluation can and cannot replace. Although CPX-RTE achieved high agreement with expert raters for many checklist-based items, residual disagreement was systematically concentrated in Patient–Physician Interaction domains that require nuanced affective and relational judgment and are only weakly inferable from dialogue transcripts. A parallel limitation applies to physical examination assessment: while agreement for physical examination items was acceptable, scoring necessarily relied on verbalized intent rather than whether the examination was performed correctly and appropriately. As a result, both interpersonal competencies and hands-on examination quality remain fundamentally underdetermined by transcript-based automation alone. These results indicate that automated OSCE evaluation should not be positioned as a fully autonomous assessor; instead, it must be embedded within a human-in-the-loop framework in which automated systems cover scalable, standardized components, while human evaluators retain responsibility for competencies that are experiential, relational, or procedural in nature.

Looking forward, learner feedback suggests that the most pressing unmet need lies not only in granular checklist scoring but also in higher-level support that helps learners *improve*, not merely *be evaluated*. Students consistently requested feedback that explains why certain questions or actions mattered, highlights missed reasoning steps, and makes progress visible across repeated encounters rather than presenting isolated, encounter-level outputs. If implemented as learners envision—combining timely feedback with clinical reasoning guidance and longitudinal progress tracking—AI-powered education tools could enable far more frequent, structured, and scalable deliberate practice than is feasible with faculty time alone. In this sense, continued advances in AI create a plausible optimistic outlook that clinical skills education could become more accessible and less dependent on local instructional resources.^25^

However, this democratizing promise should be interpreted with caution, because educational quality remains sensitive to model capability and affordability. As illustrated by our STS-based VSP evaluation, higher-capacity configurations supported more immersive and coherent simulation, whereas resource-limited configurations—though potentially “better than nothing”—were more prone to immersion-breaking conversational errors that can distract from learning. This cost–capability gradient implies an equity risk: even as AI reduces the logistical barriers to simulation-based education, settings that cannot afford sufficiently capable models (including LMICs and other resource-limited programs) may be left with systematically lower-fidelity training, thereby reproducing or widening existing disparities in educational quality. Accordingly, efforts to leverage AI for the broader availability of high-quality clinical education should explicitly define minimum fidelity thresholds and remain attentive to how model choice shapes what learners actually practice.

### Limitation

Several limitations should be considered when interpreting these findings. First, this study was conducted at a single institution and focused on a single chief complaint, requiring future multi-institutional and multi-scenario studies to enhance generalizability. Second, exposure to CPX-VSP and CPX-RTE occurred in a single session, raising the possibility of novelty effects influencing usability and immersion ratings. Third, it did not examine whether CPX-MATE translates into improvements in clinical performance. Fourth, participants were not randomly assigned to model exposure conditions, which may introduce selection or order effects. Despite these limitations, the prospective, workflow-integrated design provides a strong foundation for subsequent studies. The web-based application is currently being further developed to support ubiquitous, on-demand learning, enabling students to engage with the platform independently at any time and location, beyond formal instructional settings, while also expanding the range of clinical presentations. Ongoing work will therefore incorporate longitudinal exposure and evaluate downstream effects on CPX performance and learning transfer through randomized controlled trials.

## Conclusion

In conclusion, the development and prospective validation of CPX-MATE within an authentic CPX workflow demonstrated that CPX-VSP enables accurate and immersive VSP simulation, while CPX-RTE achieved checklist-based evaluation performance comparable to that of human faculty during HSP encounters. These findings indicate that end-to-end AI-assisted CPX platforms can be systematically evaluated across both performance and usability dimensions, supporting their potential role as complementary tools within structured CPX training while highlighting the need for careful, context-aware deployment.

## Author contributions

J.W.S.: Methodology, Software, Formal analysis, Data curation, Validation, Writing – original draft

M.K.: Software, Formal analysis, Investigation, Data curation, Validation, Writing – original draft

C.H.: Investigation, Data curation, Validation Y.S.K.: Investigation, Resources

J.C.: Investigation, Resources J.H.K.: Investigation, Resources

J.M.: Investigation, Resources A.C.: Visualization, Resources

H.Y.: Visualization, Resources S.G.W.L.: Visualization, Resources

S.C.Y.: Conceptualization, Methodology, Writing – review & editing, Supervision

C.P.: Conceptualization, Methodology, Investigation, Writing – review & editing, Project administration, Supervision

## Data Availability

The data supporting the findings of this study are not publicly available due to institutional data governance policies and the inclusion of potentially identifiable educational performance data. De-identified and aggregated data may be made available from the corresponding author upon reasonable request.

## Code Availability

The source code of the CPX-MATE platform is not publicly available due to a pending patent application and institutional intellectual property considerations. De-identified statistical analysis code supporting the findings of this study is available from the corresponding author upon reasonable request.

## Ethics approval & consent

The study was approved by the IRB of Yonsei University College of Medicine (IRB No. [4-2025-1312]). Participation was voluntary and uncompensated, and had no impact on academic evaluation. Written informed consent was obtained from all participants prior to enrollment. All recordings and transcripts were de-identified prior to analysis. The study was conducted in accordance with the Declaration of Helsinki.

## Supplementary Materials

### Method 1. Post-session usability survey instruments

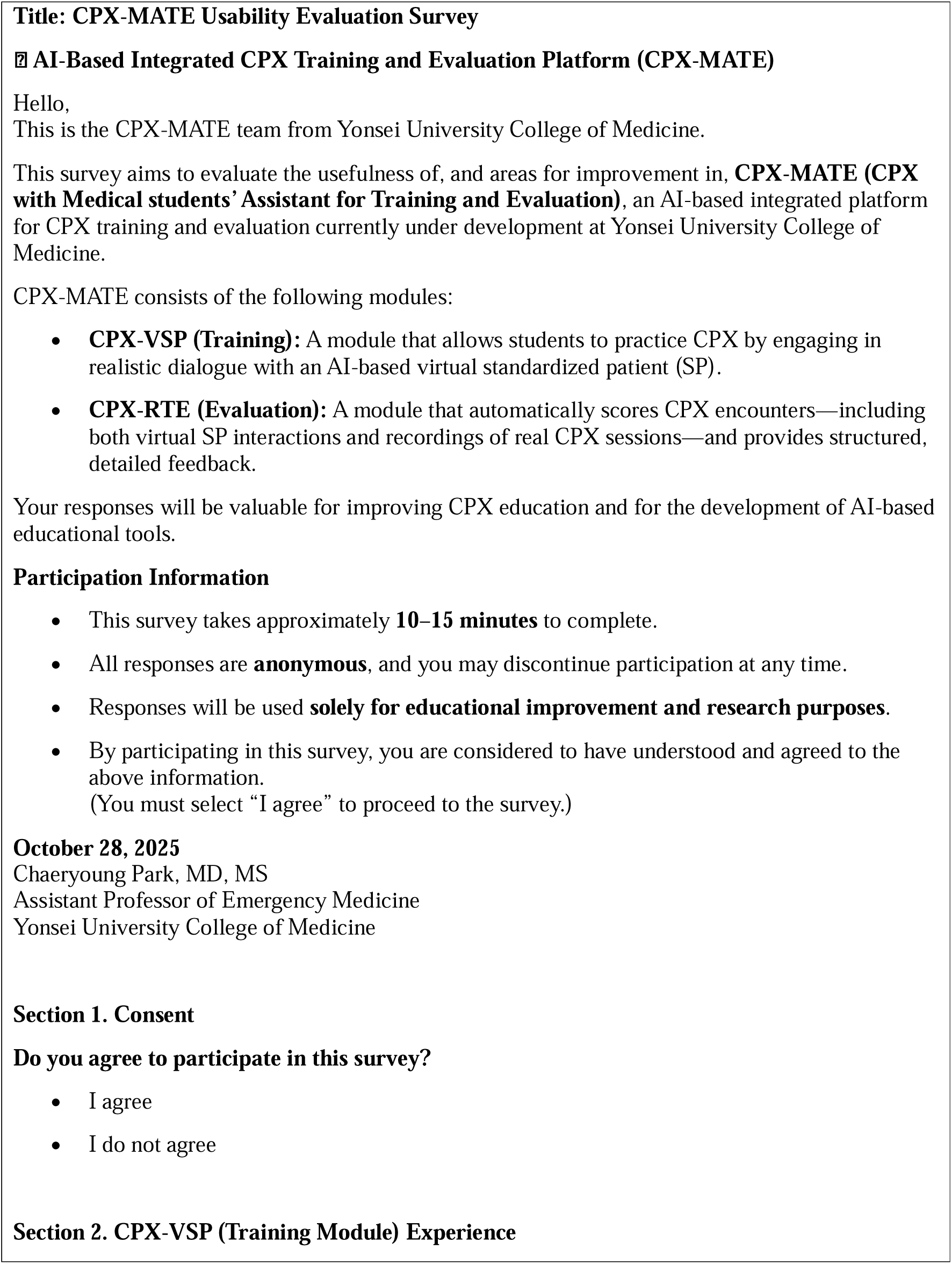
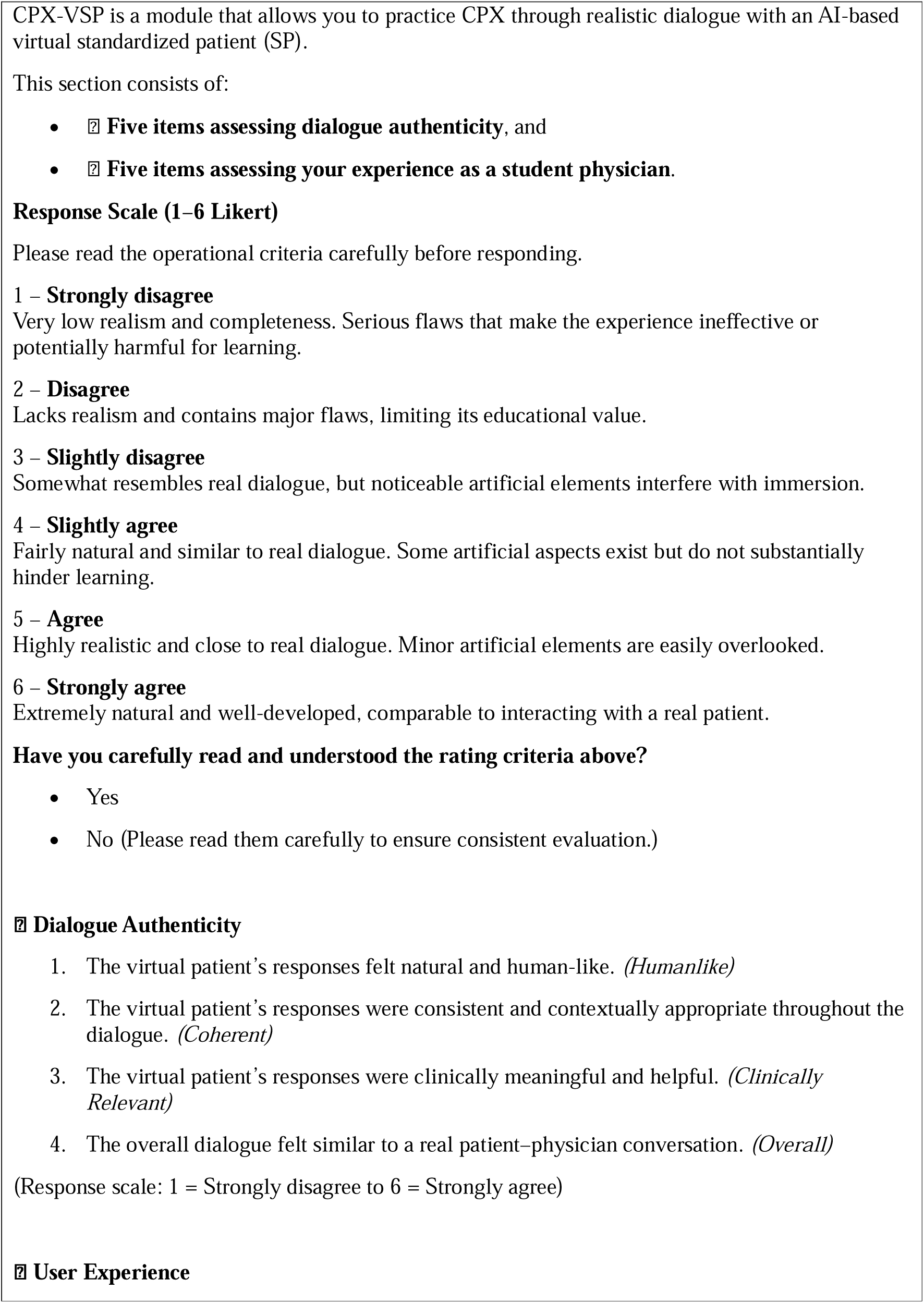
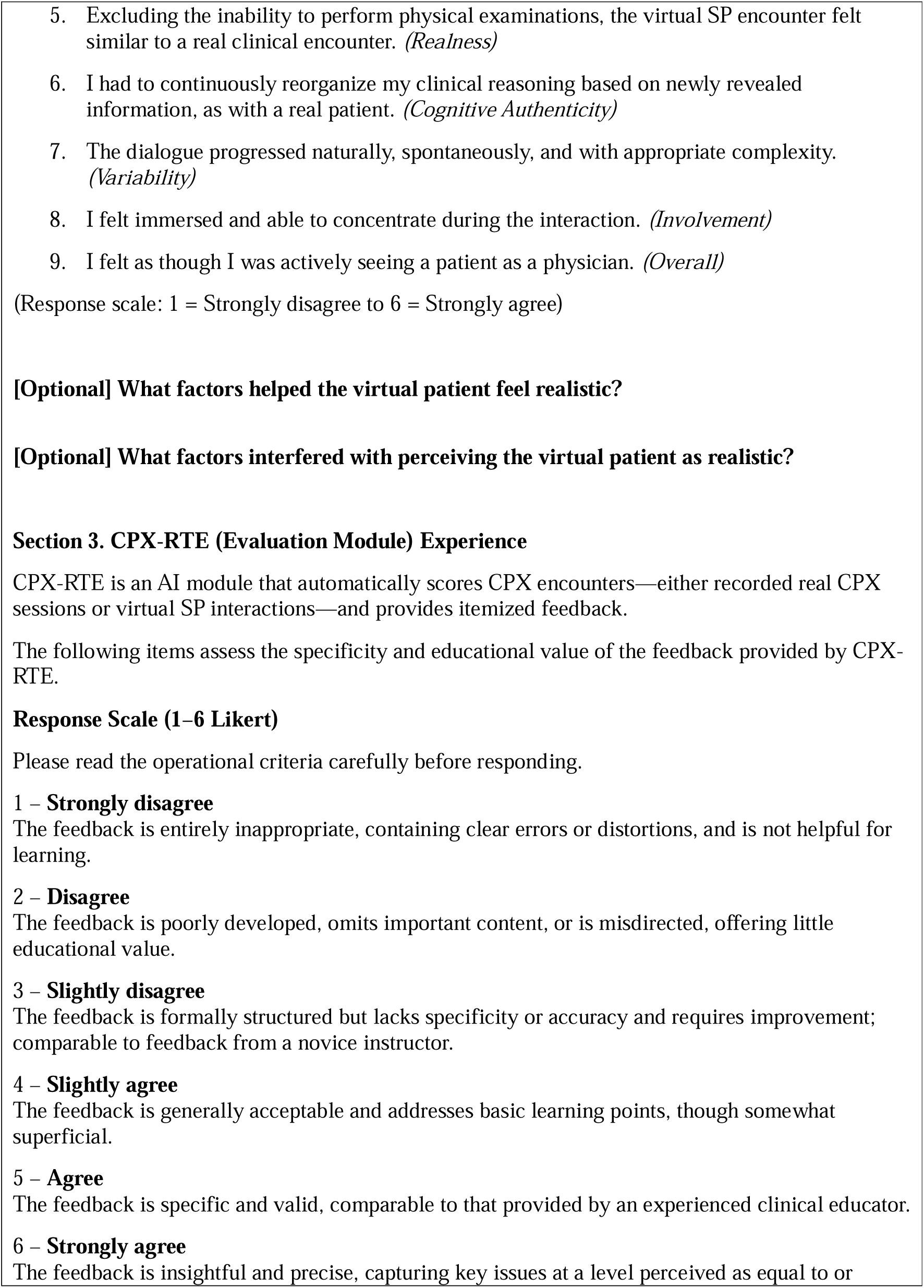
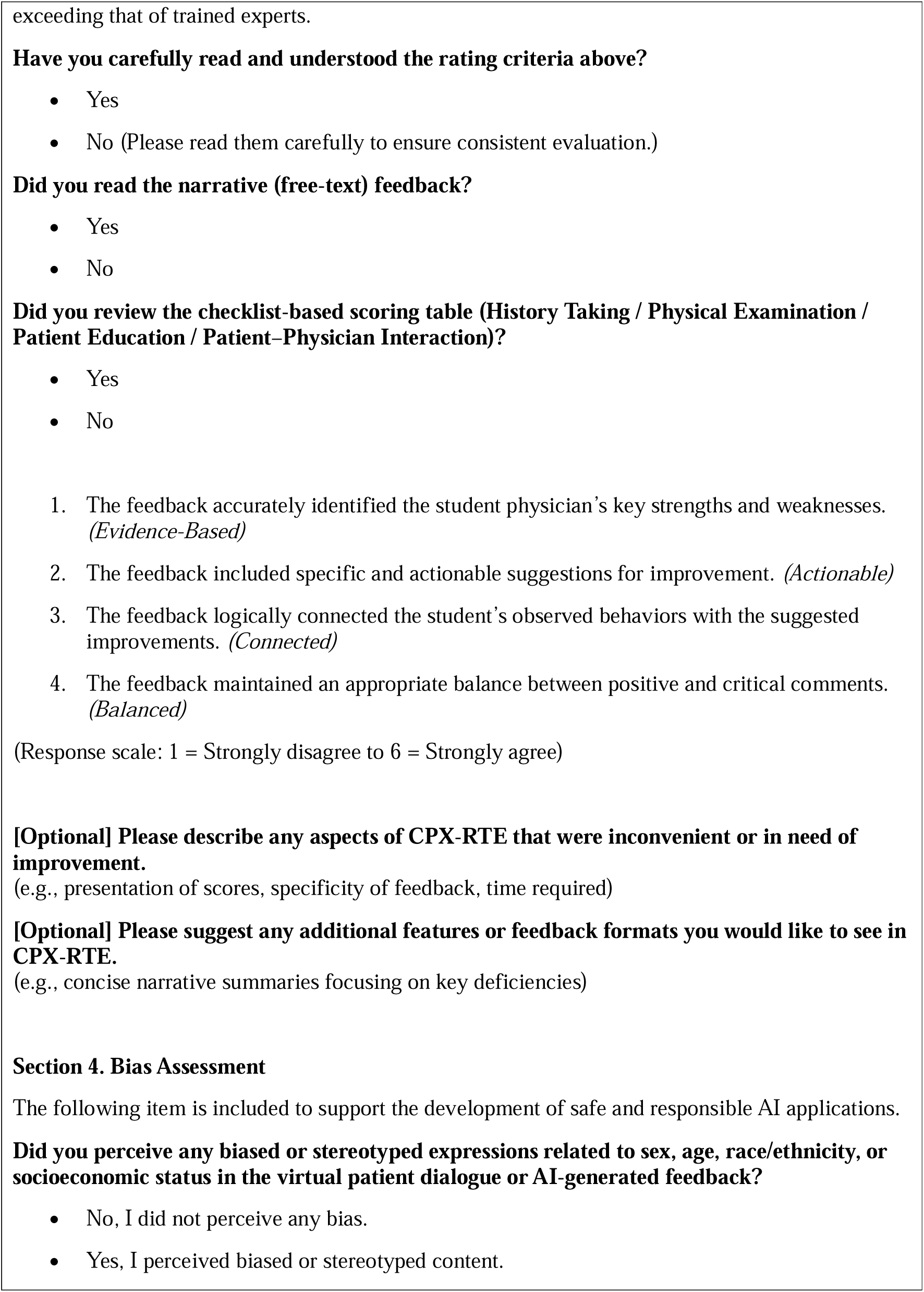
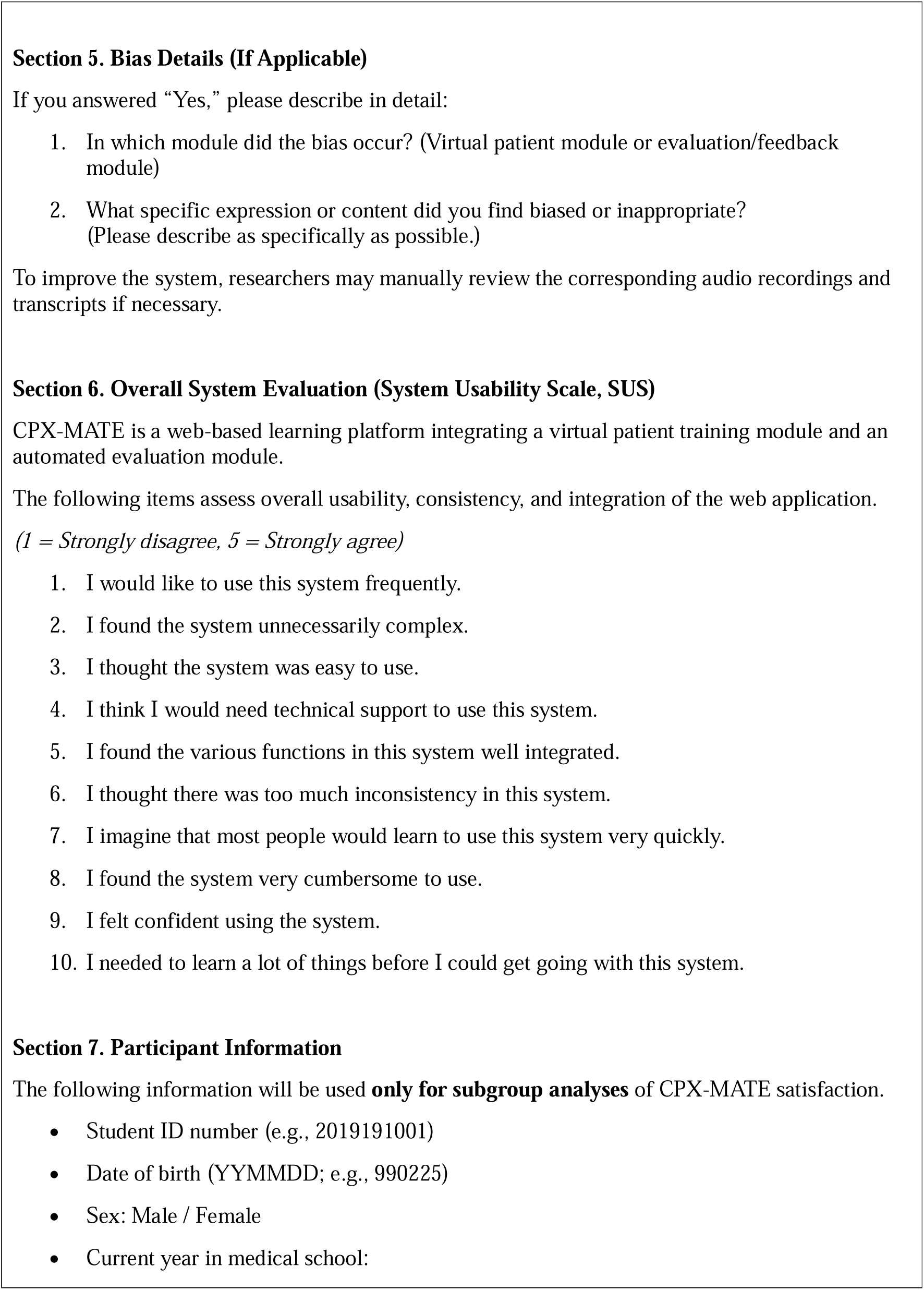
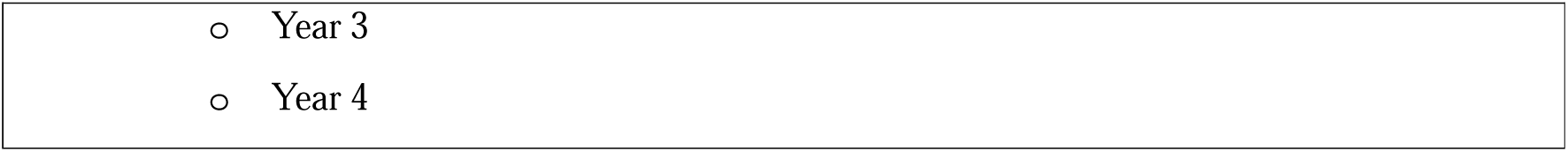

**Supplementary Table S1.** Comparison of subjective evaluation scores for CPX-VSP models (resource-limited vs full-capacity)

| Item | Resource-limited Model,<br>mean score (95% CI) | Full-capacity model,<br>mean score (95% CI) | $\Delta$ (Full-capacity - Resource-limited),<br>mean score (95% CI) | P-value |
| --- | --- | --- | --- | --- |
| Humanlike | 3.60 (3.20–3.97) | 3.93 (3.60–4.27) | 0.33 (-0.17–0.87) | 0.265 |
| Coherent | 4.03 (3.67–4.43) | 4.77 (4.43–5.13) | 0.73 (0.23–1.27) | 0.0341 |
| Clinically Relevant | 4.63 (4.20–5.03) | 4.90 (4.63–5.20) | 0.27 (-0.20–0.77) | 0.334 |
| Dialogue Overall | 3.77 (3.33–4.20) | 4.17 (3.83–4.53) | 0.40 (-0.17–0.97) | 0.235 |
| Realness | 3.57 (3.10–4.00) | 4.07 (3.73–4.40) | 0.50 (-0.00–1.03) | 0.194 |
| Cognitive Authenticity | 4.60 (4.23–4.97) | 4.73 (4.40–5.03) | 0.13 (-0.37–0.63) | 0.605 |
| Variability | 4.07 (3.67–4.47) | 4.47 (4.13–4.80) | 0.40 (-0.10–0.93) | 0.235 |
| Involvement | 3.40 (2.87–3.90) | 4.37 (4.03–4.70) | 0.97 (0.37–1.63) | 0.0188 |
| UX Overall | 3.27 (2.83–3.63) | 4.27 (3.90–4.60) | 1.00 (0.47–1.63) | 0.0100 |

Values are presented as mean (95% confidence interval). Delta represents the mean difference between models (full-capacity minus resource-limited). P values were calculated using two-sided Welch’s t-tests, with multiple comparisons adjusted using the Benjamini–Hochberg false discovery rate procedure. Confidence intervals were estimated via bootstrap resampling (1,000 iterations).

**Supplementary Table S2.** Themes Contributing to Perceived Authenticity and Immersion in CPX-VSP.

| Theme and Subtheme | Description | Illustrative quotes (English translation) | N<br>(resource-limited / full-capacity) |
| --- | --- | --- | --- |
| **1. Content-driven immersion** |  |  |  |
| **1.1 Coherent response** | Realism was driven by the perception that the VSP understood the question’s intent and responded appropriately, plausibly, and with minimal obvious errors (including | **P02:** “The responses were quite appropriate... Even when I interrupted, it continued without issues.” | **8 / 10** |
|  | preparedness for minor questions). | **P06:** “It felt prepared even for minor questions.”<br>**P15:** “It expressed the clinical presentation well.”<br>**P54:** “Even with the cushioning language typical in CPX, it grasped the point and answered.” |  |
| **2. Interactional/affective realism** |  |  |  |
| **2.1 Patient-like voice** | Natural, everyday speech and a patient-like tone (sometimes resembling a real SP) enhanced authenticity. | **P23:** “The speech felt natural—like talking to a real person.” **P55:** “The tone sounded like a patient / a real person.”<br>**P04:** “It answered the way real patients would.” | **4 / 6** |
| **2.2 Affective and reactive** | Emotional expression and “reactions” (adding small extra remarks) were perceived as human-like behavioral cues. | **P22:** “Natural reactions, and it added extra remarks.”<br>**P53:** “Emotional acting.” | **0 / 2** |
| **2.3 Responsive dialogue flow** | Smooth turn-taking (including after interruption), natural conversational flow, and handling unexpected turns/questions supported perceived realism. | **P02:** “Even when I interrupted, it continued without issues.”<br>**P42:** “The fact that it was a real dialogue made it feel realistic.”<br>**P49:** “It felt everyday and like a real patient; the flow followed my questions naturally.”<br>**P34:** “It included an unexpected question mid-conversation.” | **3 / 2** |

**Supplementary Table S3.** Themes Contributing to Loss of Immersion and Interactional Limitations.

| Theme and Subtheme | Description | Illustrative quotes (English translation) | N<br>(resource-limited / full-capacity) |
| --- | --- | --- | --- |
| **1. Content breaks clinical realism** |  |  |  |
| **1.1 Tangential responses** | Immersion was disrupted when the VSP misunderstood the intent of questions, produced answers unrelated to the asked domain, or responded with clinically incongruent content that broke the conversational logic. | **P10:** “When I said I would examine the lungs, it answered about an abdominal exam—it felt scripted and broke immersion.”<br>**P33:** “At times it gave answers that didn’t match my question.” | **12 / 3** |
| **1.2 Scripted system-like** | Role-play broke when the VSP abruptly used system-like phrasing or stepped out of the patient role (e.g., rigid template phrases or “not in the scenario” responses). | **P04:** “A stiff phrase like ‘It is normal’ suddenly appeared and broke immersion.”<br>**P04:** “When asked about past jobs, it replied, ‘That isn’t in the scenario.’” | **2 / 0** |
| **1.3 Oversharing** | Participants reported reduced realism when the VSP volunteered information before being asked, provided overly complete/organized details, or answered “too helpfully,” which felt unlike real patient communication and distorted time management. | **P50:** “It felt overly helpful; it often tells everything before I even ask, which is unlike real patients.”<br>**P29:** “I only asked about alcohol/smoking, but it volunteered frequency and amount on its own.” | **9 / 2** |
| **1.4 Repetition** | Realism was reduced when the VSP repeated the same answer across slightly different questions, making the exchange feel patterned or mechanical. | **P06:** “With slightly different questions, it often repeated the same answer multiple times.” | **1 / 0** |
| **2. Interactional/affective realism** |  |  |  |
| **2.1 Monotone / not in pain** | Participants noted immersion loss when voice qualities (tone, prosody, intonation) felt “AI-like,” monotone, or incongruent with a | **P09:** “The TTS sounded lively rather than like a sick patient, so immersion was difficult.” | **8 / 4** |
|  | patient in pain (e.g., overly bright/energetic voice). | **P56:** “It had the typical AI-like speaking style and intonation.” |  |
| **2.2 Limited empathy cues** | Participants described difficulty maintaining a patient–physician relationship when empathy/rapport cues were sparse or when suffering/pain was not conveyed (verbally or nonverbally), making the interaction feel less human. | **P18:** “Because it felt like talking to a machine, I also ended up questioning mechanically rather than building rapport.”<br>**P47:** “Without nonverbal cues about pain/state, it was hard to insert empathy appropriately.” | **3 / 3** |
| **3. System & modality constraints** |  |  |  |
| **3.1 Latency** | Delays between question and answer (time lag) disrupted the rhythm of CPX-style interaction and reduced realism. | **P07:** “There was a delay in answering, which made it feel less real.”<br>**P53:** “With long questions/explanations, the time lag became very noticeable.” | **4 / 6** |
| **3.2 Dialogue breakdown** | Immersion broke when responses intermittently stopped, conversations were cut off, or the system failed mid-dialogue. | **P32:** “After I explained the abdominal CT, it suddenly stopped responding and the dialogue broke.” | **0 / 4** |
| **3.3 Physical exam unavailability** | Participants reported reduced realism due to constraints intrinsic to the modality: physical exams being performed “by description,” inability to observe the patient’s appearance, and lack of embodied clinical cues. | **P05:** “Not seeing the patient, and having to do the physical exam verbally, broke immersion.”<br>**P21:** “Not being able to perform the physical exam.” | **5 / 5** |
| **3.4 Screen-mediated awkwardness** | The screen-mediated setting itself (not facing a person) was experienced as awkward and less authentic, independent of content quality. | **P39:** “It felt awkward because I wasn’t speaking face-to-face.”<br>**P42:** “I ended up staring at the screen while talking, which felt less like a real encounter.” | **0 / 2** |

**Supplementary Table S4.**
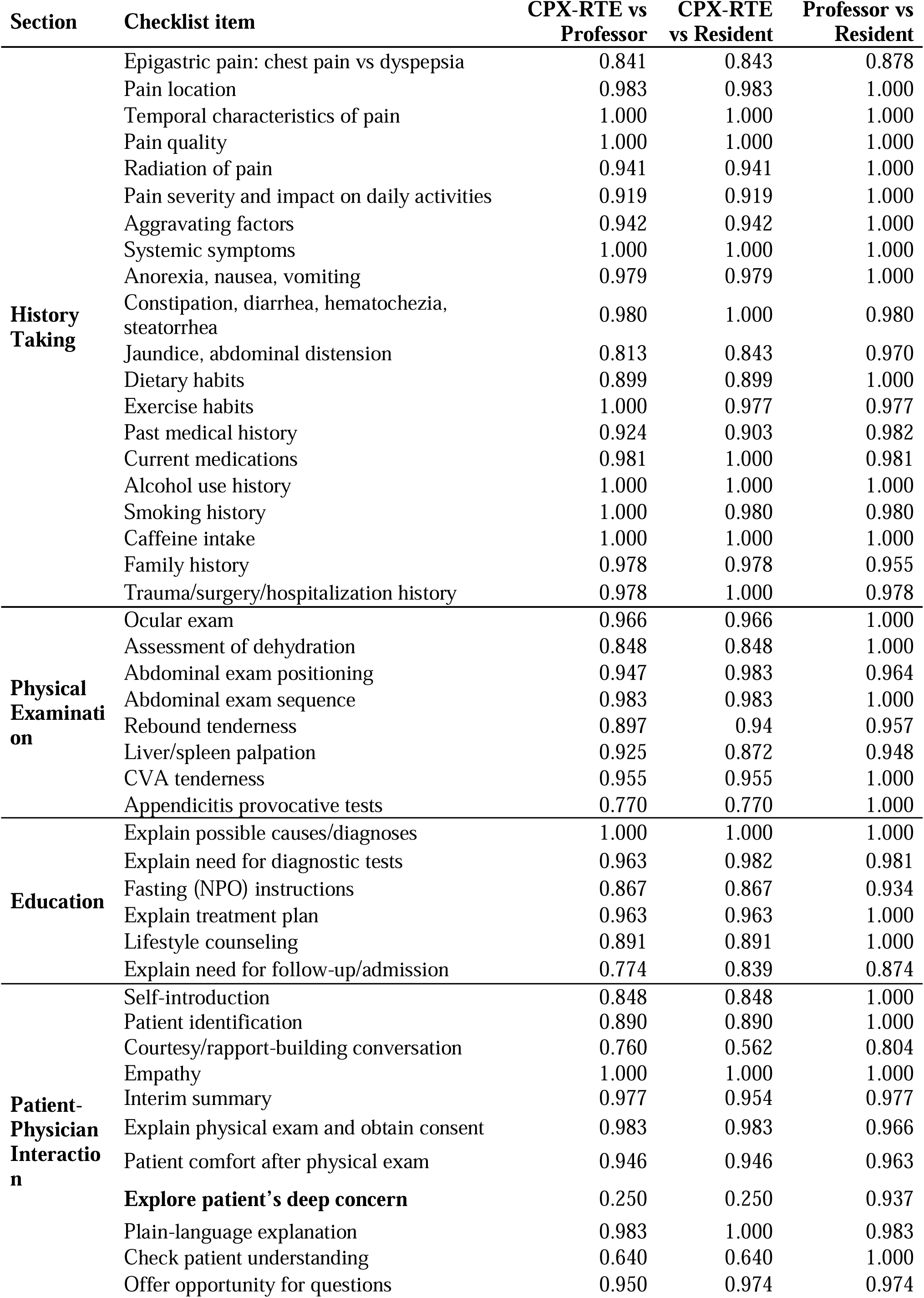
CPX-RTE item-based evaluation (Gwet’s AC1)

**Supplementary Table S5.** Themes Related to Scoring Accuracy Concerns and Perceived Educational Utility of CPX-RTE.

| Theme and Subtheme | Description | Illustrative quotes (English translation) | n (all) |
| --- | --- | --- | --- |
| **1. Structured feedback - checklist content** |  |  |  |
| **1.1 Strict or non-contextual** | Learners felt the checklist sometimes applied rigid criteria that did not reflect clinical context (e.g., time constraints, symptom patterns), leading to perceived unfair deductions or “forced” differentials. | **P04:** “I skipped radiation pain due to time... but the scoring seems to apply strict criteria without considering context.”<br>**P04:** “The system makes us cover even unlikely differentials, which feels unrealistic.” | 6 |
| **1.2 Need for prioritization** | Participants suggested adding item weights or priority levels (e.g., points per item) and surfacing “most important missing items” first. | **P02:** “If you could show weights or points per checklist item, that would help.”<br>**P11:** “It would help if missing items were displayed first as a priority.” | 4 |
| **1.3 Granular empathy items** | Some requested more granular evaluation of relational/communication elements, such as empathy and rapport, beyond biomedical checklist items. | **P07:** “Evaluation of rapport/empathy could be more detailed.” | 2 |
| **2. Structured feedback - scoring** |  |  |  |
| **2.1 Scoring errors in CPX-RTE** | A common concern was misclassification of performed actions (e.g., “I did it” marked as “not done”), undermining trust in the scoring. | **P18:** “I verified the patient, but it was recorded as not done.”<br>**P31:** “Toward the end, my voice wasn’t recognized and my score dropped.” | 2 |
| **3. Feedback/evaluation UI/UX** |  |  |  |
| **3.1 Readability and priority** | Learners found narrative feedback lengthy or mixed with praise, making key improvement points hard to spot; they wanted concise summaries and prioritized issues. | **P09:** “Praise is nice, but improvements don’t stand out.”<br>**P26:** “Please summarize only what needs correction and highlight priorities.” | 13 |
| **3.2 Detailed guidance** | Many requested more concrete, actionable guidance—example questions/phrases for missing items, “what to ask next,” and brief rationales tied to differential diagnosis. | **P10:** “For items marked incorrect, include what question I should have asked.”<br>**P20:** “Include example phrases, not just ‘ask about X.’”<br>**P23:** “Adding rationales (why each question/exam matters) would<br>help learning.” | 15 |
| **3.3 Feedback format** | Users requested more structured or customizable presentation (e.g., table-based scoring, clearer structured feedback, removing emojis). | **P02:** “Remove emojis.”<br>**P28:** “If the scoring were presented more intuitively in a table, it would be better.” | 6 |
| **3.4 Time management support** | Some suggested time-support features (optional timer, end-of-session alerts, section-wise time tracking) to better train CPX pacing. | **P12:** “Show time spent on history/physical/education to help improve pacing.”<br>**P19:** “Instead of a visible timer, an alert at 2 minutes remaining could work.” | 4 |
| **3.5 Tracking across sessions** | Learners wanted longitudinal summaries of recurring gaps across repeated sessions and benchmarking (e.g., peer averages). | **P26:** “Across repeated cases, summarize what I repeatedly miss.”<br>**P35:** “It would be helpful to compare with others’ average scores.” | 3 |

## References

1 Ba, H., Zhang, L., He, X. & Li, S. Knowledge mapping and global trends in the field of the objective structured clinical examination: bibliometric and visual analysis (2004-2023). JMIR Medical Education 10, e57772 (2024).

2 Dewan, P., Khalil, S. & Gupta, P. Objective structured clinical examination for teaching and assessment: Evidence-based critique. Clinical Epidemiology and Global Health 25, 101477 (2024).

3. Lavigne, E., et al. AI-Standardized Clinical Examination training on OSCE performance. NEJM AI 2, AIoa2500066 (2025).

4 Patrício, M. F., Julião, M., Fareleira, F. & Carneiro, A. V. Is the OSCE a feasible tool to assess competencies in undergraduate medical education? Medical teacher 35, 503–514 (2013).

5 Mukurunge, E., Nyoni, C. N. & Hugo, L. Assessment approaches in undergraduate health professions education: towards the development of feasible assessment approaches for low-resource settings. BMC Medical Education 24, 318 (2024).

6 Ataro, G., Worku, S. & Asaminew, T. Experience and challenges of objective structured clinical examination (OSCE): perspective of students and examiners in a clinical department of Ethiopian University. Ethiopian Journal of Health Sciences 30 (2020).

7 Cook, D. A., Overgaard, J., Pankratz, V. S., Del Fiol, G. & Aakre, C. A. Virtual patients using large language models: Scalable, contextualized simulation of clinician-patient dialogue with feedback. Journal of Medical Internet Research 27, e68486 (2025).

8 Holderried, F., et al. A language model–powered simulated patient with automated feedback for history taking: Prospective study. JMIR Medical Education 10, e59213 (2024).

9 Yamamoto, A., et al. Enhancing medical interview skills through AI-Simulated patient interactions: nonrandomized controlled trial. JMIR medical education 10, e58753 (2024).

10 Luo, M.-J. et al. A large language model digital patient system enhances ophthalmology history taking skills. NPJ Digital Medicine 8, 502 (2025).

11 Jacobs, C. et al. Application of AI Communication Training Tools in Medical Undergraduate Education: Mixed Methods Feasibility Study Within a Primary Care Context. JMIR Med Educ 11, e70766 (2025). 10.2196/70766

12. Pu, Y. et al. in Proc. Interspeech 2025. 26-30.

13 Takata, T., Yamada, R., René, A. O. N., Xu, K. & Fujimoto, M. in 2024 Joint 13th International Conference on Soft Computing and Intelligent Systems and 25th International Symposium on Advanced Intelligent Systems (SCIS&ISIS). 1–5 (IEEE).

14. OpenAI. Introducing gpt-realtime, <https://openai.com/ko-KR/index/introducing-gpt-realtime/> (2025).

15. Google. Get started with Live API, <https://ai.google.dev/gemini-api/docs/live?hl=ko&example=mic-stream> (2025).

16 Lee, Y. C. Rethinking artificial intelligence in medicine: from tools to agents. Clinical and experimental emergency medicine 12, 101 (2025).

17. Geathers, J. et al. in International Conference on Artificial Intelligence in Education. 231-245 (Springer).

18 Shakur, A. H., et al. Large Language Models for Medical OSCE Assessment: A Novel Approach to Transcript Analysis. arXiv preprint arXiv:2410.12858 (2024).

19 Holcomb, M. J. et al. Zero-Shot Multimodal Question Answering for Assessment of Medical Student OSCE Physical Exam Videos. medRxiv, 2024.2006. 2005.24308467 (2024).

20. OpenAI. Introducing Whisper, <https://openai.com/ko-KR/index/whisper/> (2025).

21. OpenAI. Introducing GPT-5, <https://openai.com/ko-KR/index/introducing-gpt-5/> (2025).

22 Roh, H. et al. [Development of guide to clinical performance and basic clinical skills for medical students]. Korean J Med Educ 27, 309–319 (2015). 10.3946/kjme.2015.27.4.309

23 Lewis, J. R. The system usability scale: past, present, and future. International Journal of Human–Computer Interaction 34, 577–590 (2018).

24 Ahmed, S. K. et al. Using thematic analysis in qualitative research. Journal of Medicine, Surgery, and Public Health 6, 100198 (2025).

25. Khan, S. Sal Khan’s 2023 TED Talk: AI in the classroom can transform education, <https://blog.khanacademy.org/sal-khans-2023-ted-talk-ai-in-the-classroom-can-transform-education/> (2023).

